# Smartwatch-based loss-of-pulse detection for out-of-hospital cardiac arrest: the BECA-S2 and BECA-S6 studies

**DOI:** 10.64898/2026.11.25.26368935

**Authors:** Roelof G. Hup, Maud E. Kortman, Robin W.F.H. Kuijpers, Michelle J.S. Mensinck, Chaimae Bouchnaf, Alexandre J. Ouss, Hanno L. Tan, Reinder Haakma, Lukas R.C. Dekker, Xi Long, Rik Vullings

## Abstract

**Objective:** Out-of-hospital cardiac arrest (OHCA) is often unwitnessed, which delays treatment and lowers the chance of survival. A smartwatch could act as a witness by detecting loss of pulse from wrist photoplethysmography (PPG) and subsequently alert emergency services. We aimed to develop and evaluate such a detection model.

**Approach:** As smartwatch data of OHCA are extremely difficult to acquire, we used in-hospital proxy data to test sensitivity: wrist PPG and accelerometry recorded during baseline and induced loss-of-pulse periods in 31 patients during catheter ablation of ventricular arrhythmias and defibrillation threshold (DFT) testing during subcutaneous implantable cardioverter-defibrillator (S-ICD) implantation (BECA-S2). In total, 33 loss-of-pulse events, annotated using accompanying electrocardiograms and, where available, arterial blood pressure, were included. To count false alarms, we used wrist PPG and accelerometry of 100 healthy volunteers during unsupervised daily activities (BECA-S6). A recurrent neural network was evaluated with cross-validation, with one fold per patient with events. As the electrical VF induction and defibrillation shocks applied during these procedures would not be present in true OHCA episodes, we tested for and mitigated the resulting target leakage.

**Main results:** The Area Under the Receiver Operating Characteristic Curve (AUC-ROC) was 86.8% (95% CI: 79.7 to 92.5%) and the Area Under the Precision-Recall Curve (PR-AUC) was 52.4% (95% CI: 38.6 to 66.2%). At a false alarm rate of zero on BECA-S6, the event-level sensitivity on BECA-S2 was 48.5% (95% CI: 32.1 to 65.5%). Higher sensitivities came at the cost of more false alarms. Without mitigation, target leakage inflated the model scores by 35.3% (95% CI: 11.5 to 61.4%) of the model’s discriminative range.

**Significance:** These results show the feasibility of smartwatch-based loss-of-pulse detection. Larger studies with data more representative of OHCA are needed. We recommend that future studies on induced loss of pulse test for and mitigate target leakage.

## 1 Introduction

Out-of-hospital cardiac arrest (OHCA) is one of the leading causes of mortality worldwide, with an incidence of emergency medical services-treated OHCA of 35.7–100.0 per 100,000 population per year and survival to hospital discharge rates of only 3.4% to 15.8% [1, 2]. Successful treatment of OHCA consists of early recognition, alerting emergency medical services (EMS), starting cardiopulmonary resuscitation (CPR), and early application of advanced life support, together known as the *chain of survival*.

A large proportion (34.4–55.1% according to national registries) of OHCA cases are unwitnessed [1]. Given that victims are unable to call for help themselves, this delays the first step of the chain of survival and therefore the whole treatment. As survival rates of OHCA drop rapidly with delayed treatment (≈ 6% per minute), the chances of survival in unwitnessed cases are poor (*<*5%) [3, 4].

In the BEating Cardiac Arrest (BECA) project, our aim is to develop a smartwatch-based solution that functions as a witness when no human witness is present [5]. We use wrist-based photoplethysmography (PPG) and accelerometry to detect loss of pulse and loss of motion, the main symptoms of OHCA, which would enable starting the chain of survival in unwitnessed cases.

Several research groups and manufacturers have shown interest in this concept in parallel, converging on largely the same problem statement but differing in device, algorithm, and validation approach. Google Research has implemented a loss-of-pulse feature on the Pixel Watch 3, combining digital signal processing with a convolutional neural network across a three-stage detection cascade [6]. In prospective validation, it reached a sensitivity of 67.2% in arterial-occlusion testing, with 1.34 algorithm-level false positives (check-in notifications) per user-year in free-living conditions.

The DETECT research group has validated a rule-based wrist-PPG algorithm in patients with induced and spontaneous circulatory arrest during cardiac procedures, reporting sensitivities of 92–98% at 0.12–0.26 false alarms per hour [7, 8]. The HEART-SAFE consortium has taken a device-independent approach using consumer smartwatches, validating detection across cuff-induced ischemia, cardioplegic cardiac arrest during surgery, and animal models, reaching 90.3% sensitivity but a comparatively modest 94.1% hour-level specificity, and has additionally demonstrated end-to-end alerting through a community-responder network [9]; a subsequent case report documented the first automated detection of a genuine spontaneous cardiac arrest by a consumer smartwatch [10].

Although these studies show cautiously promising sensitivity, specificity in real-world conditions remains the central open problem, as frequent false alarms would burden emergency medical services and community responders, and could lead users to stop wearing the device or to ignore its alarms. Reported false alarm rates vary by orders of magnitude across studies and validation settings [6, 7, 8, 9], evidence on spontaneous (as opposed to induced) cardiac arrest remains limited to small cohorts and case reports [7, 10, 11, 12], and the authors of most studies call for further validation before real-world deployment.

In this work, we take a step toward such validation by presenting a methodology for the detection of loss of pulse using wrist-PPG and accelerometry and evaluating it on both clinical and real-world data, which, to our knowledge, has not been done before. As smartwatch data of true OHCA events are extremely difficult to obtain in view of their low incidence [1, 2], we approximated such data with two studies. We tested detection sensitivity based on in-hospital measurements during routine cardiac procedures in which ventricular tachycardia or fibrillation (VT/VF) was expected to occur (the BECA-S2 study), and false alarm rate in out-of-hospital measurements in healthy volunteers wearing the device in unsupervised daily life (the BECA-S6 study). As electrical VF induction and defibrillation shocks that start or terminate some of these events leave signal artifacts that would not be present in OHCA, a model could exploit these artifacts instead of detecting the loss of pulse itself (target leakage). Therefore, unlike previous studies, we tested for and mitigated target leakage.

## 2 Methods

### 2.1 The BECA-S2 study of intraoperative loss of pulse

To measure detection sensitivity, the prospective observational BECA-S2 study included patients who underwent catheter ablation of ventricular arrhythmias or defibrillation threshold (DFT) testing during subcutaneous implantable cardioverter-defibrillator (S-ICD) implantation between August 2023 and February 2026 at Catharina Hospital (Eindhoven, The Netherlands). These procedures were chosen because of the high probability that VT/VF events would occur. Patients were eligible when they were at least 18 years old, able to provide informed consent, and had no wounds, injuries, infectious diseases, or tattoos on the wrist on which the PPG/accelerometer detection device was placed. The cohort consisted of the patients who presented for these procedures and met these criteria.

During the procedures, data were acquired from the catheterization lab (cathlab) using the GE HealthCare CardioLab EP and Mac-Lab Recording Systems. This included ECG (sampled at 240-977 Hz) and, if available, arterial blood pressure (ABP, sampled at 977 Hz). Furthermore, patients wore a Philips Elan data logger wristband, equipped with a single-channel green light photoplethysmography (PPG) sensor sampling at 32 Hz and a three-axis accelerometer sampling at 128 Hz. For redundancy purposes, patients wore a wristband on each wrist if the procedure allowed for it.

After data collection, the onsets and ends of VT/VF events were independently annotated in the cathlab ECG and ABP data by two electrophysiologists, followed by adjudication of disagreements by consensus-seeking. Episodes of VF visible in the ECG were considered to cause loss of pulse.

Episodes of VT were considered to cause loss of pulse only if ABP was available and showed a peak arterial pressure of 50 mmHg or lower. Based on these annotations, we calculated inter-annotator agreement using Gwet’s AC1 and Cohen’s kappa, and the annotation overlap using the median Intersection over Union (IoU). An expert in photoplethysmography translated these annotations to the Philips Elan data, annotating the onsets and ends of loss of pulse in the PPG signals.

Pre- and postoperative parts of the wristband signals were excluded, leaving only the intraoperative data. Given the precarious nature of the cardiovascular system during catheter ablation, we only included the wristband data of these procedures that were accompanied by ECG and/or ABP signals to confirm both events and absence of events. Since ECG and ABP were not recorded continuously, this split each record into several segments. As the S-ICD implantation procedures were generally uneventful, we included all of their intraoperative wristband data, including parts without ECG or ABP in which the absence of events could not be confirmed.

The records were cut after each occurrence of loss of pulse, after which 10 seconds of data were excluded to allow the cardiovascular system to recover. In some procedures, periodic blood pressure measurements were performed using an automated oscillometric sphygmomanometer, leading to arterial occlusions in the arm that mimic loss of pulse. These episodes were excluded with a 10-second margin to avoid confusing the detection model. Loss-of-pulse episodes shorter than 3 seconds were excluded with a 10-second margin as well. After all cuts, only segments of 30 seconds or longer were included. Loss-of-pulse events that fell within excluded data were not analysed.

### 2.2 The BECA-S6 study of out-of-hospital free-living conditions

To measure the false alarm rate, we performed the prospective observational BECA-S6 study, in which we collected wrist PPG and accelerometry of healthy volunteers without loss of pulse.

Between July 2023 and April 2024, participants were recruited from a pool of university students, employees, and community volunteers. Eligibility required a minimum age of 18 years, absence of wrist wounds, injuries, and tattoos, and unrestricted upper-limb mobility. Participants wore two Philips Elan data logger wristbands, one on each wrist, for approximately one hour in free-living conditions.

### 2.3 Preprocessing and feature extraction

After data acquisition, the PPG signals were corrected for hardware-controlled amplitude scaling and separated into baseline and pulsatile components by third-order causal low-pass and high-pass Butterworth filters with a cutoff frequency of 0.5 Hz. Accelerometry signals were downsampled by a factor of 4 to match the PPG sampling frequency, and their gravity/DC component was removed by a third-order causal high-pass Butterworth filter with a cutoff frequency of 0.5 Hz.

We extracted 37 time- and frequency-domain features per sample, including PPG derivatives, PPG moments (variance, skewness, kurtosis), the Hjorth parameters [13], zero-crossing rate, peak-to-peak amplitude and its log-ratio against the same quantity 10, 30, 60, and 120 seconds earlier, signal energy, signal-to-noise ratio, spectral entropy and flux, acceleration and jerk magnitude, peak-to-peak acceleration amplitude log-sum and log-ratios against delayed amplitudes, and acceleration moments. Where applicable, the features are calculated using a sliding window of 2 seconds (time-domain) or 8 seconds (frequency-domain). A complete description of the features is provided in the Appendix A.

After feature extraction, all features are z-score normalized using a causal exponential moving average of mean and variance, updated every second from past data in that record. The averaging window retained an effective memory of approximately 20 minutes. To allow the normalization and the longest feature window to settle, the detector was armed only after a record had 2 minutes of history and each segment 10 seconds of data; earlier seconds were not scored.

Some VT/VF events were started by electrical VF induction and/or terminated by a defibrillation shock, both of which provide a clear signal marker in both PPG and accelerometry that would otherwise not be present in OHCA. As these markers coincide with the onset or end of loss of pulse, a model could learn to recognize these markers instead of the loss of pulse itself, which would inflate its performance on these data but not carry over to OHCA. This is a form of target leakage: information that reveals the label during development but is not available in practice. To avoid this, we opted to gap these signal parts by overwriting the normalized features with the running average. Furthermore, we added a binary indicator feature that marks which seconds were gapped, informing the detection model which parts to handle differently. As the gaps themselves still coincide with these events, the detection model is additionally discouraged from using information from the gaps, as described in the next subsection.

### 2.4 Detection model

The detection model consists of a recurrent neural network with a convolutional encoder. The shared-weights encoder takes one second of features as input and works it through three convolutional blocks before global average pooling. The encoder outputs of consecutive seconds are then used as input to a long short-term memory (LSTM) network, followed by a shared-weights decoder with a dense layer to make the final logit classification of loss of pulse. Model width and encoder dropout were explored as hyperparameters during model selection (see Section 2.5). An overview of the model is shown in Figure 1 and Appendix B.

**Figure 1:**
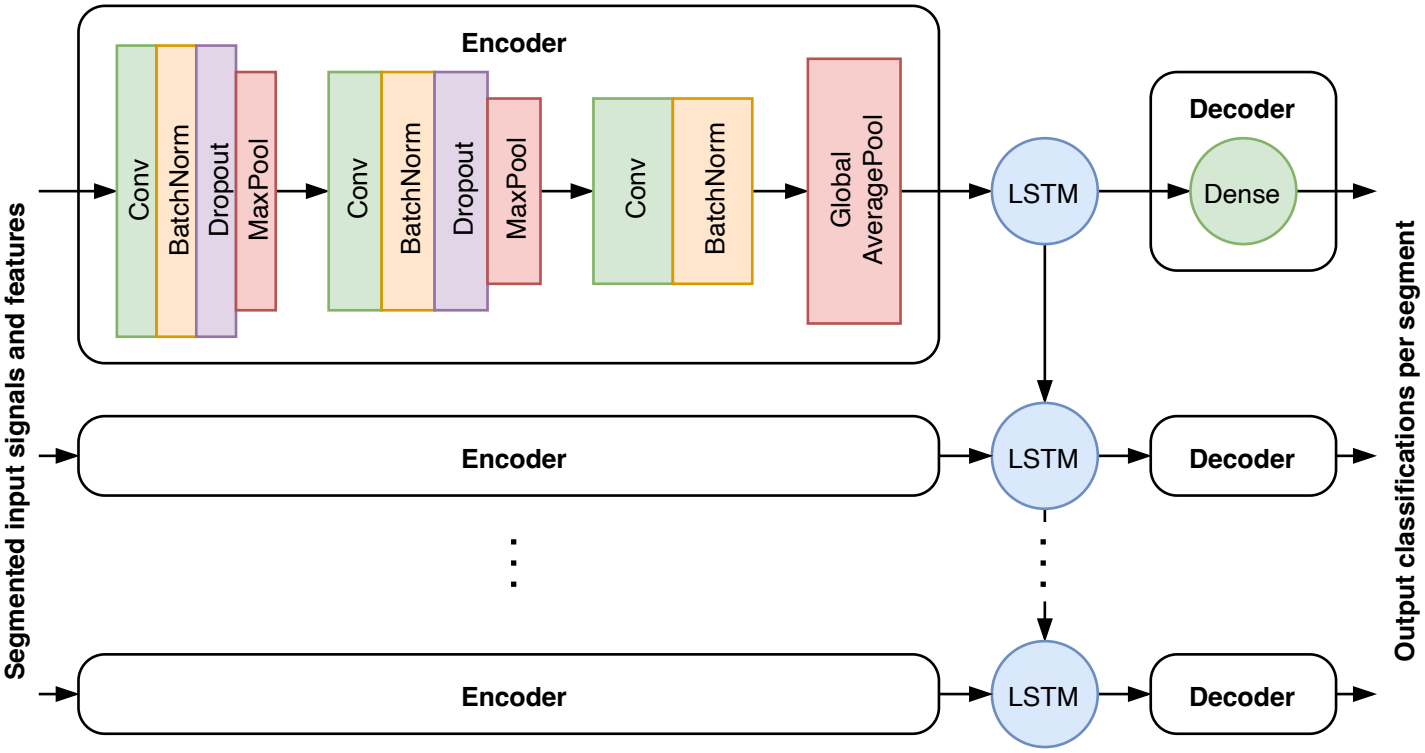
Overview of the recurrent neural network detection model, consisting of a shared-weights encoder, a long short-term memory network, and a shared-weights decoder.

This detection model was trained using an Adam optimizer (*lr* = 0.001) on Tversky loss [14]. The soft Tversky counts are

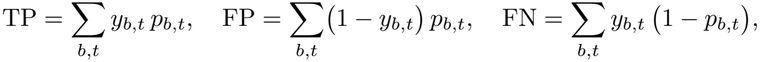

where *y∈* [0, 1]^*B×T*^ are the per-second targets in a batch, and *p ∈*[0, 1]^*B×T*^ the predicted per-second probabilities.

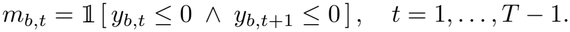

Furthermore, we applied a smoothness penalty penalizing noisy consecutive estimates. For the smoothness function, we define a mask defined only between adjacent non-positive-labeled seconds to avoid penalizing genuine event onsets. The penalty itself is the mean squared error of consecutive estimates over this mask.

In addition, we applied a gradient penalty GP(*z*), the squared gradient of the output logits *z* with respect to all input channels of the gapped input samples, to penalize reliance on the gapped input and thus mitigate target leakage [15]. The full loss function is then

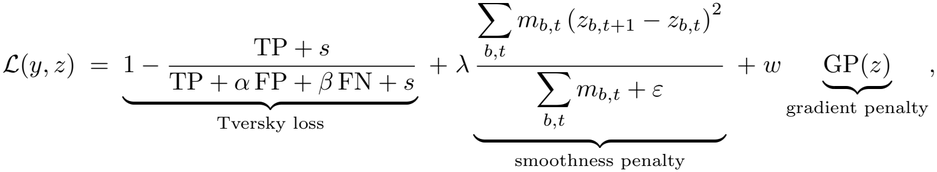

with smoothing constant *s* = 1 and *ε* = 10^*−*6^. The Tversky weights *α, β* (false-positive and false-negative weights, respectively), the smoothness weight *λ*, and the gradient-penalty weight *w* are hyperparameters. The model was fed with batches of 32 arbitrary-length (30-60 seconds) pieces of signal and features, where half of the batch was guaranteed to contain loss-of-pulse-positive seconds. Online amplitude jittering of ±5% was applied.

As a device should raise alarms for events rather than for individual seconds, the second-level classifications were converted into event-level alarms. To prevent incidental positive classifications from triggering a false alarm, we used a debouncing mechanism, requiring a certain number of consecutive positive second-level classifications before an alarm was raised. The required number of positive classifications (the debounce length, 1 to 60 seconds) was chosen per fold after model selection (see Section 2.5).

### 2.5 Evaluation

We used cross-validation to assess detection performance, with one fold for each BECA-S2 patient having at least one loss-of-pulse event. The BECA-S2 patients without events and the BECA-S6 participants (with data from both wrists) were randomly and uniformly distributed over the folds, forming one group per fold. In each fold, one group was used for testing, the next five groups for validation, and the remaining groups for training. Inside each fold, the model hyperparameters (model width, encoder dropout, Tversky loss parameters, smoothness penalty weight) were selected using successive halving on the Area Under the Precision-Recall curve (PR-AUC), starting with 27 Latin-Hypercube-sampled hyperparameter candidates trained for 5 epochs, of which the 9 best were trained for 15 epochs. The 3 best of these were selected as finalists and trained until convergence on validation loss, and the finalist with the highest validation PR-AUC was selected. As hyperparameters were selected within each fold, the reported performance reflects the training procedure rather than a single fixed model. The optimal debounce length was chosen per fold on the validation data. For each candidate length, we determined the event-level false alarm rate (FAR) needed to reach 80% sensitivity, with sensitivity determined on the BECA-S2 validation patients and false alarms counted on the BECA-S6 validation participants. We then selected the length with the lowest 97.5th-percentile upper bound of this FAR, obtained from a cluster bootstrap of 1,000 replicates. Only debounce lengths up to the median duration of the validation events were considered, so that at least half of these events lasted long enough to be detected.

To mitigate target leakage, the training procedure was repeated for all gradient-penalty weights in {0, 1, 10, 10^2^, 10^3^, 10^4^, 10^5^, 10^6^, 10^7^}. The hyperparameter search was performed at weight 0. To reduce computational effort, for all other weights, each fold reused the configuration selected for that fold at weight 0.

To determine whether target leakage remained, we probed each trained model with synthetic gaps: if a model relies on gaps as a cue for loss of pulse, inserting a gap raises its scores. For each weight, residual target leakage was estimated by inserting synthetic gaps, copied from real gaps, in event-free data of the test BECA-S2 patients and BECA-S6 participants (arrest-free context; 10 gaps per data segment) and just before loss-of-pulse events (arrest context; 10 gaps per event) and observing the change in model scores in the 30 seconds after the synthetic gap. Only events not already preceded by a gap were eligible for the arrest-context probe. This leakage effect was expressed relative to the discriminative range of the model, i.e., the difference between the mean model score during loss-of-pulse events and during event-free data. For both contexts, we then tested whether the leakage effect lay within a pre-specified tolerance of ±5% of this range (an equivalence test), using a bootstrap over patients and participants (20,000 replicates).

The gradient-penalty weight was then selected as follows. Weights showing model collapse, i.e., with a discriminative range of less than 25% of that of the unmitigated model (*w* = 0), were excluded from weight selection. Subsequently, fixed-sequence testing was applied to find the smallest gradient-penalty weight that, together with all larger non-collapsed weights, passed both tolerance tests: weights were tested one at a time from the largest non-collapsed weight downward, each at a significance level of 0.05, stopping at the first weight failing either test. As the tests are performed in a fixed order, no correction for multiple testing is needed. The PR-AUC of the selected weight was compared with that of the unmitigated model (*w* = 0) by a paired patient-cluster bootstrap (20,000 replicates) of the difference in macro-averaged PR-AUC over the test patients.

For each selected model, the second-level classifications on the test patient were generated, from which the Receiver Operating Characteristic (ROC) and PR curves were inferred by threshold-sweeping. A 500-point grid was used to interpolate the curves of each patient before macro-averaging across patients. Patients without loss-of-pulse events have no defined curves and do not contribute to the average. We inferred 95% confidence intervals using patient-cluster bootstrap with 2,000 replicates, resampling all patients. The same procedure was used to determine Area Under the ROC curve (AUC-ROC) and PR-AUC.

We also tested event-level performance, where an event-level alarm was defined as a sequence of second-level positives reaching the debounce length and was considered a true positive whenever its onset was within a detection window of 10 seconds before a loss of pulse onset or 30 seconds after it, reflecting that a device alarms on events rather than individual seconds. Sensitivity was determined on the BECA-S2 events, and false alarms were counted on BECA-S6 data. The false alarm rate is the number of alarms divided by the number of scored hours, pooled over the BECA-S6 participants in a test fold. As each wrist was recorded and scored separately, an hour of recording on both wrists counts as two wrist-hours. In a secondary analysis, false alarms were also counted on BECA-S2, to compare false alarm rates between clinical and real-world conditions.

After applying debouncing on the test patients, the resulting Free-response ROC (FROC) curves were weighted based on event count (micro-averaging) to mitigate the low count of events per patient. Bootstrapping over cross-validation folds (each holding one BECA-S2 patient with events and the BECA-S6 participants of that fold) was used to infer 95% confidence intervals. Each FROC curve was summarized by four pre-specified points: the FAR at sensitivities of 60%, 80%, and 95%, and the sensitivity at a FAR of zero.

Lastly, we examined whether the algorithm’s performance differed by participant characteristics, separately for each cohort. In BECA-S2, we examined whether per-patient PR-AUC differed by procedure type (catheter ablation vs. DFT testing), sex (male versus female), age (decades), BMI (kg/m^2^), Fitzpatrick skin tone (ordinal), or wrist circumference (cm). Because PR-AUC is bounded between 0% and 100%, we used fractional-logit regression (binomial family, logit link) [16] with all six characteristics entered simultaneously. Effects are reported as average marginal effects in percentage points. In BECA-S6, we examined whether the number of false alarms per participant differed by sex, age (decades), BMI, Fitzpatrick skin tone, or wrist circumference (averaged over both wrists). We used a Poisson regression of the false alarm count of both wrists summed, with the logarithm of the monitored hours as an offset and all five characteristics entered simultaneously. False alarms were counted at the threshold and debounce length that each cross-validation fold selected for 80% validation sensitivity. Effects are reported as average marginal effects in false alarms per hour. For both models, confidence intervals and two-sided *p*-values were obtained by bootstrapping, resampling patients in BECA-S2 and cross-validation folds in BECA-S6 (as participants within a fold share a threshold), with Bonferroni correction per cohort (six and five comparisons, respectively; *α* = 0.05). Collinearity was assessed with generalized variance inflation factors (GVIF), with GVIF *<* 5 considered safe [17].

### 2.6 Ethics statement

An ethics exemption was granted by the Medical Ethics Review Board (METC) of Máxima MC, Veldhoven, for BECA-S2 (5 December 2022, N22.082) and BECA-S6 (19 June 2023, N23.054), and BECA-S6 was additionally approved by the Ethical Review Board of Eindhoven University of Technology (27 June 2023, ERB2023EE5). Written informed consent was obtained from all patients and participants. Both studies were carried out according to the Declaration of Helsinki and local statutory requirements.

## 3 Results

### 3.1 Analysis datasets

The BECA-S2 study recruited 32 patients, for whom 42 events of loss of pulse were annotated. The annotators showed substantial agreement [18] (Cohen’s *κ*of 0.722), and Gwet’s AC1, which is less affected by the low prevalence of loss of pulse, was 0.999. The overlap between the annotations is captured by a median Intersection over Union of 0.891. After data preparation, the analysis population consists of 31 patients with 33 loss-of-pulse events and 31 hours of data: one patient had no monitored interval of 30 seconds or longer, and 9 events were excluded because they lasted shorter than 3 seconds (6), occurred in monitored intervals shorter than 30 seconds (2), or lacked ECG/ABP coverage (1). An example of an annotated loss-of-pulse event, preceded by electrical VF induction, is shown in Figure 2.

**Figure 2:**
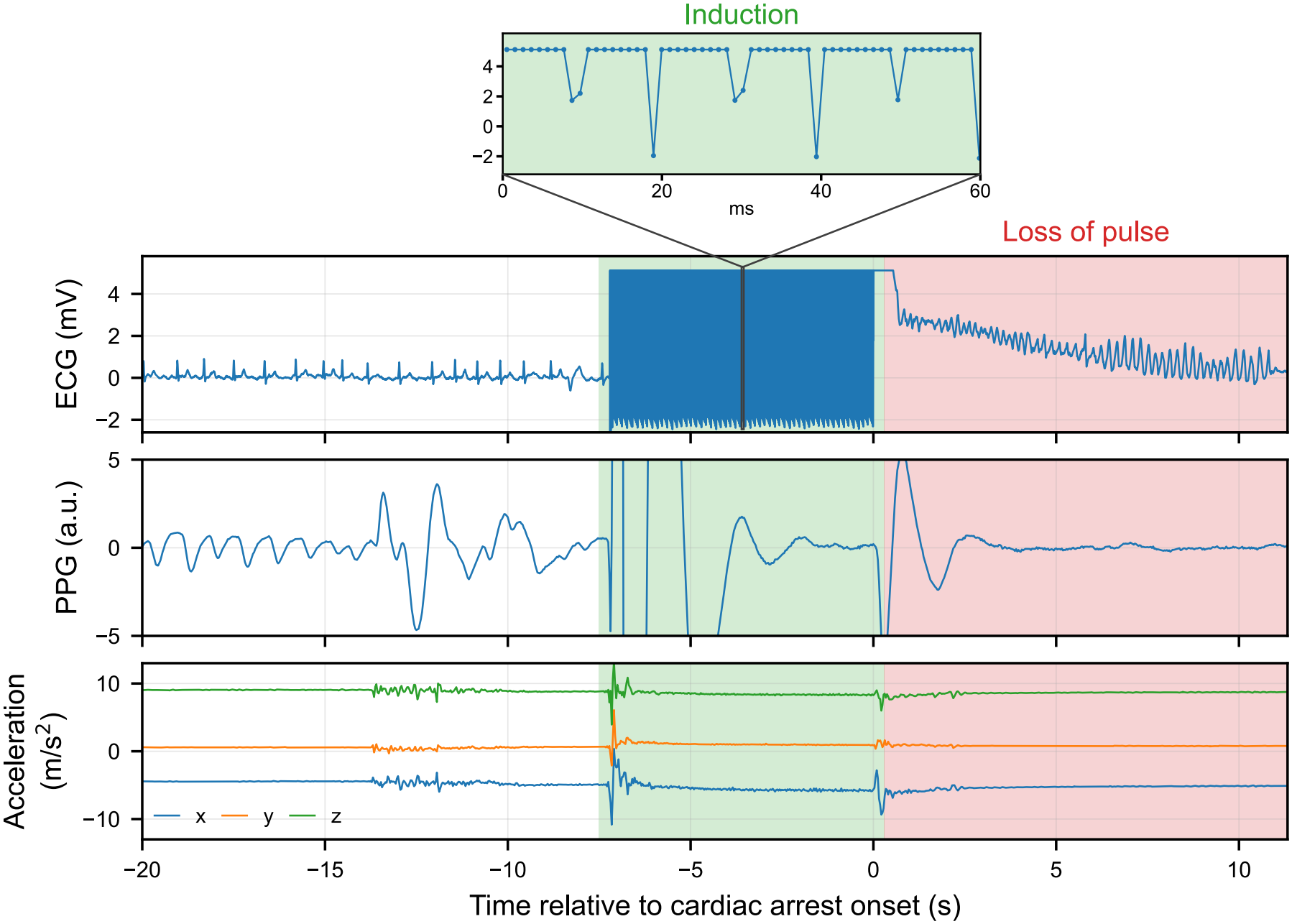
Example of a loss-of-pulse event preceded by electrical VF induction, with the electro-cardiogram, photoplethysmogram, and three-axis accelerometry signals shown relative to the onset of cardiac arrest. The induction (green) produces a clear artifact in the photoplethysmogram and accelerometry signals that would not be present in out-of-hospital cardiac arrest, and was therefore gapped before model input. The annotated loss of pulse (red) starts at the end of the induction. No arterial blood pressure was available for this event.

The BECA-S6 study recruited 102 participants, of whom 2 were excluded due to general health concerns and limited mobility, leaving 100 participants in the analysis population contributing 156.7 wrist-hours (median of 1.59 wrist-hours per participant). The population flowcharts and patient and participant characteristics for both studies can be found in Figure 3 and Table 1, respectively. The BECA-S2 patients were predominantly male (80.6%) and had light skin tones (Fitzpatrick type I or II in 27 of the 28 patients with a recorded skin tone), whereas the BECA-S6 participants were more balanced in sex (56.0% male) and mostly had Fitzpatrick type III (75.0%).

**Table 1:**
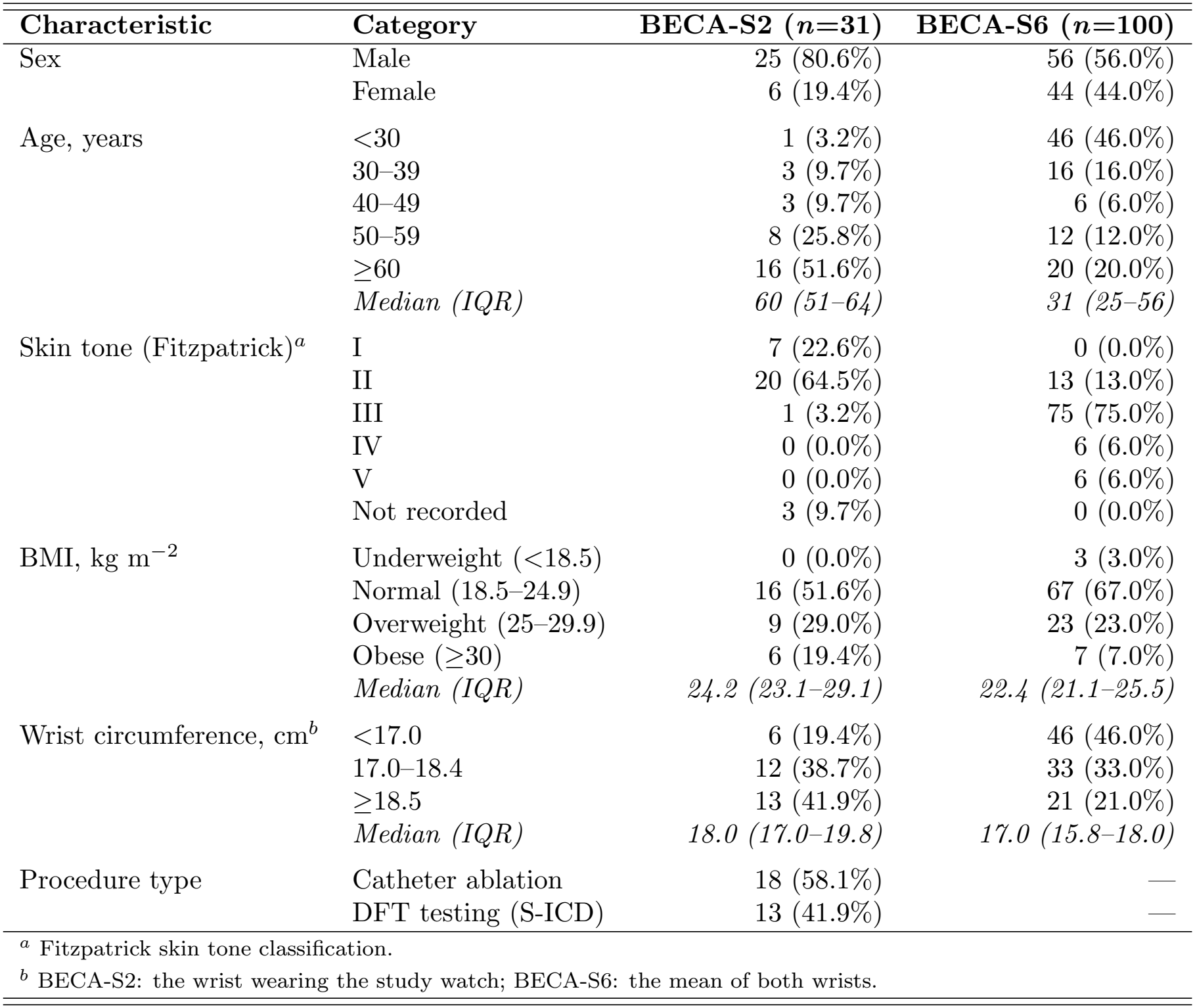
Characteristics of the BECA-S2 patients and BECA-S6 participants. Values are *n* (%) unless indicated otherwise.

| **Characteristic** | **Category** | **BECA-S2 (*n*=31)** | **BECA-S6 (*n*=100)** |
|---|---|---|---|
| Sex | Male | 25 (80.6%) | 56 (56.0%) |
|  | Female | 6 (19.4%) | 44 (44.0%) |
| Age, years | <30 | 1 (3.2%) | 46 (46.0%) |
|  | 30–39 | 3 (9.7%) | 16 (16.0%) |
|  | 40–49 | 3 (9.7%) | 6 (6.0%) |
|  | 50–59 | 8 (25.8%) | 12 (12.0%) |
|  | ≥60 | 16 (51.6%) | 20 (20.0%) |
|  | *Median (IQR)* | *60 (51–64)* | *31 (25–56)* |
| Skin tone (Fitzpatrick)<sup>*a*</sup> | I | 7 (22.6%) | 0 (0.0%) |
|  | II | 20 (64.5%) | 13 (13.0%) |
|  | III | 1 (3.2%) | 75 (75.0%) |
|  | IV | 0 (0.0%) | 6 (6.0%) |
|  | V | 0 (0.0%) | 6 (6.0%) |
|  | Not recorded | 3 (9.7%) | 0 (0.0%) |
| BMI, kg m<sup>−2</sup> | Underweight (<18.5) | 0 (0.0%) | 3 (3.0%) |
|  | Normal (18.5–24.9) | 16 (51.6%) | 67 (67.0%) |
|  | Overweight (25–29.9) | 9 (29.0%) | 23 (23.0%) |
|  | Obese (≥30) | 6 (19.4%) | 7 (7.0%) |
|  | *Median (IQR)* | *24.2 (23.1–29.1)* | *22.4 (21.1–25.5)* |
| Wrist circumference, cm<sup>*b*</sup> | <17.0 | 6 (19.4%) | 46 (46.0%) |
|  | 17.0–18.4 | 12 (38.7%) | 33 (33.0%) |
|  | ≥18.5 | 13 (41.9%) | 21 (21.0%) |
|  | *Median (IQR)* | *18.0 (17.0–19.8)* | *17.0 (15.8–18.0)* |
| Procedure type | Catheter ablation | 18 (58.1%) | — |
|  | DFT testing (S-ICD) | 13 (41.9%) | — |

<sup>*a*</sup> Fitzpatrick skin tone classification.
<sup>*b*</sup> BECA-S2: the wrist wearing the study watch; BECA-S6: the mean of both wrists.

**Figure 3:**
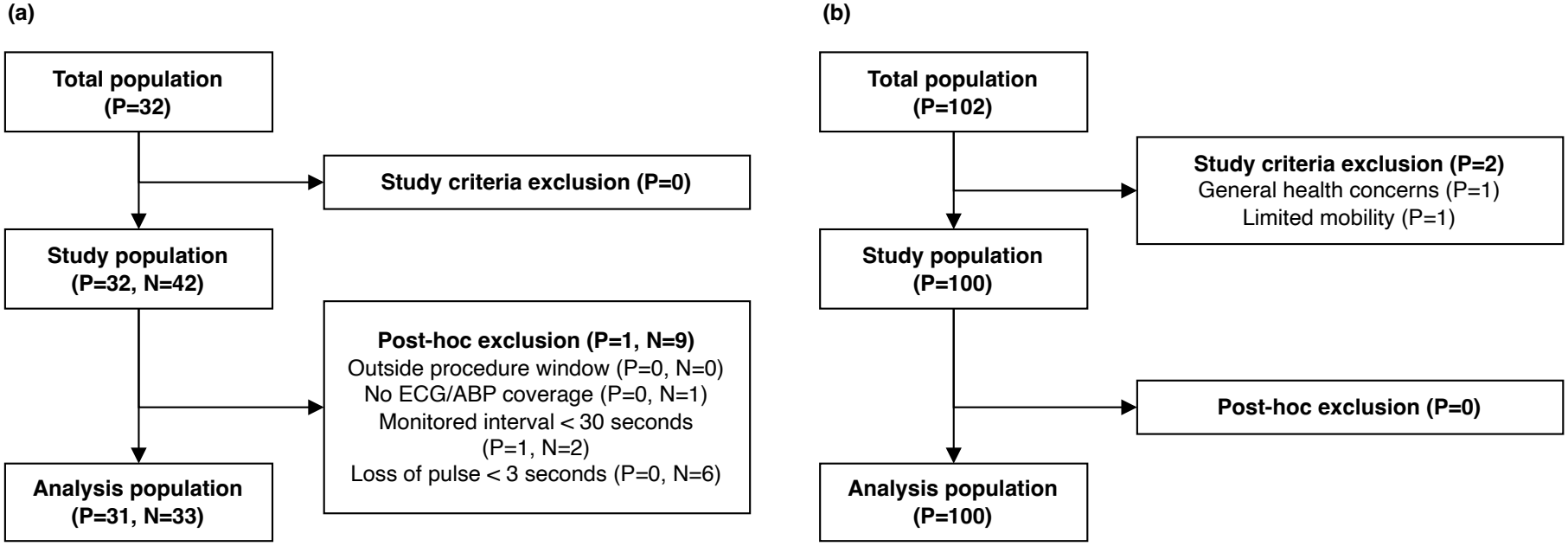
Population flowcharts of (a) the BECA-S2 study and (b) the BECA-S6 study. *P* indicates the number of patients (BECA-S2) or participants (BECA-S6), *N* the number of loss of pulse events.

The 22 BECA-S2 patients with events were used to define a 22-fold cross-validation procedure, over which the 9 BECA-S2 patients without events and the 100 BECA-S6 participants were distributed. In each fold, 16 BECA-S2 patients with events (plus 3–9 without events) and 70–76 BECA-S6 participants were used for training, 5 patients with events (plus 0–5 without events) and 20–25 participants for validation, and 1 patient with events (plus 0–1 without events) and 4–5 participants for testing.

### 3.2 Target leakage mitigation analysis

Of the 33 events, 13 events in 7 patients were not preceded by a gap and were therefore eligible for the arrest-context probe. With no target leakage mitigation, i.e., a gradient-penalty weight of *w* = 0, injecting a synthetic gap in the arrest context shifted the model’s per-second score by a mean of 4.273 logit units (*n* = 7), corresponding to 35.3% (95% CI: 11.5 to 61.4%) of the discriminative range. In the arrest-free context, the shift in score was 0.809 logit units or 6.69% (95% CI: 4.78 to 9.88%) of the discriminative range (*n* = 131 patients and participants). The tolerance tests for both contexts could not show that these shifts fall within the 5% tolerance (*p*_tol_ = 0.990 and *p*_tol_ = 0.955, respectively), indicating the need for mitigation.

As the models at gradient-penalty weights 10^6^ and 10^7^ showed discriminative ranges (1.43 and 0.17 logit units) below 25% of that of the unmitigated model (12.09 logit units), we considered these models as collapsed, excluding them from the weight selection. For the remaining weights, we observed a decrease in target leakage effects in both arrest- and arrest-free contexts with increasing weight, indicating that the gradient penalty affects target leakage (see Figure 4(a) and (b)).

**Figure 4:**
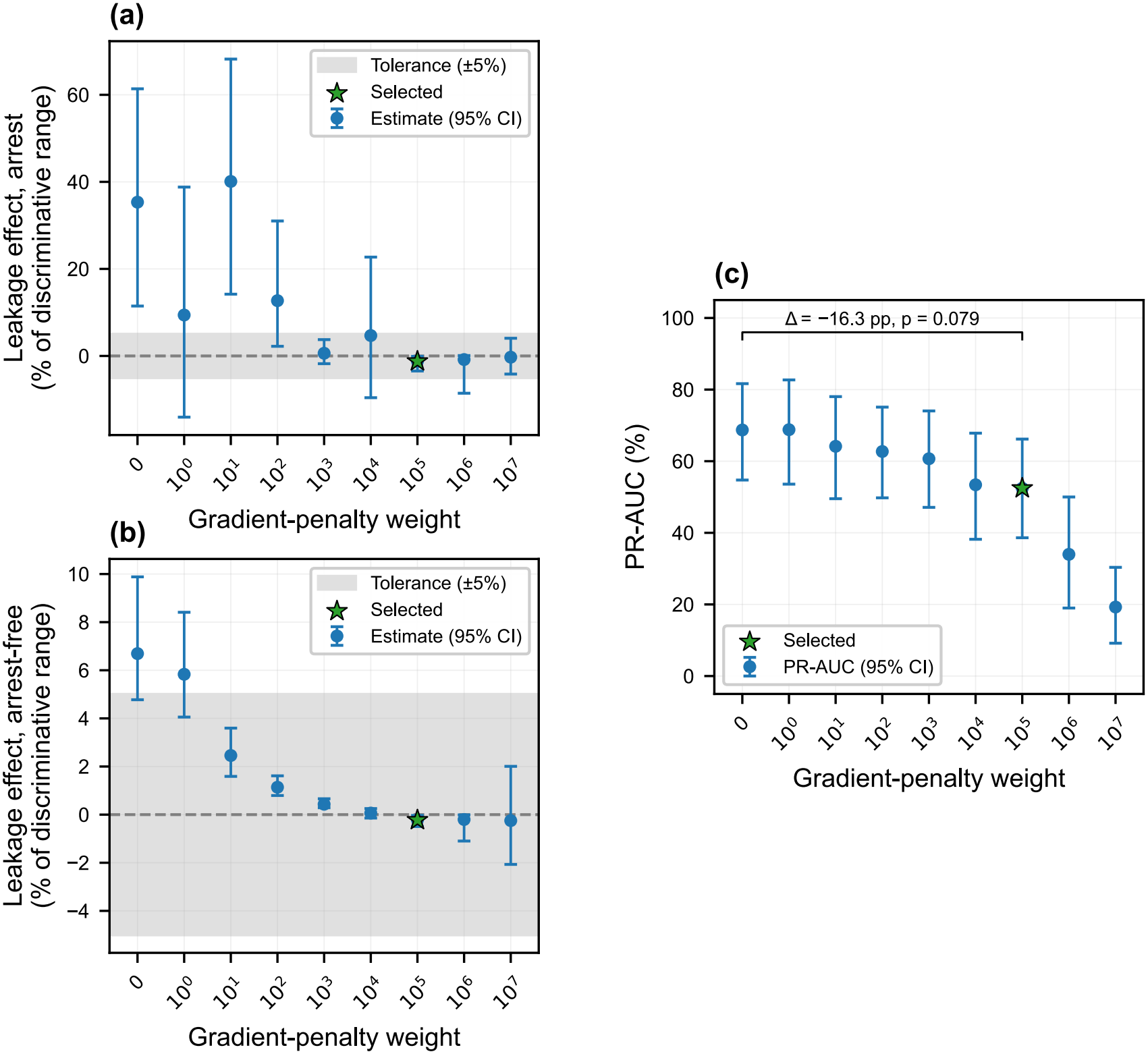
(a) Target leakage effect in the 30 seconds following a synthetic gap in the arrest context vs. the 5% discriminative range tolerance for different gradient-penalty weights. (b) Target leakage effect in the 30 seconds following a synthetic gap in the arrest-free context (BECA-S2 and BECA-S6 combined) vs. the 5% discriminative range tolerance for different gradient-penalty weights. (c) Area under the precision-recall curve on the test patients for different gradient-penalty weights. The bracket gives the paired difference in this area between the selected weight and weight 0.

The subsequent fixed-sequence testing led to the adoption of 10^5^, which passed both tolerance tests, whereas the next weight in the sequence (10^4^) failed the arrest-context test and ended the sequence. At 10^5^, we observed an arrest-context shift in score of −0.045 logit units, corresponding to −1.23% (95% CI: −3.46 to −0.09%) of the discriminative range. In an arrest-free context, the mean shift in score is −0.008 or −0.23% (95% CI: −0.49 to −0.03%) of the discriminative range. Both fall within the tolerance (*p*_tol_ = 0.003 and *p*_tol_ *<* 0.001, respectively). The reduction in leakage was accompanied by a lower test-set PR-AUC point estimate, from 68.7% (95% CI: 54.7 to 81.7%) at gradient-penalty weight 0 to 52.4% (95% CI: 38.6 to 66.2%) at weight 10^5^ (see Figure 4(c)). The paired difference of −16.3 pp (95% CI: −34.1 to 1.8) was not statistically significant (two-sided paired bootstrap, *p* = 0.079).

### 3.3 Classification performance

The classification model achieved a second-level AUC-ROC of 86.8% (95% CI: 79.7 to 92.5%) and PR-AUC of 52.4% (95% CI: 38.6 to 66.2%), showing overall good classification performance, with wide confidence intervals reflecting relatively large inter-patient variability. The curves were averaged over the 22 patients with loss-of-pulse events. For reference, a random classifier has a precision equal to the loss-of-pulse prevalence, which was 0.536% when pooled over these patients. The corresponding ROC and PR curves are shown in Figure 5(a) and (b).

**Figure 5:**
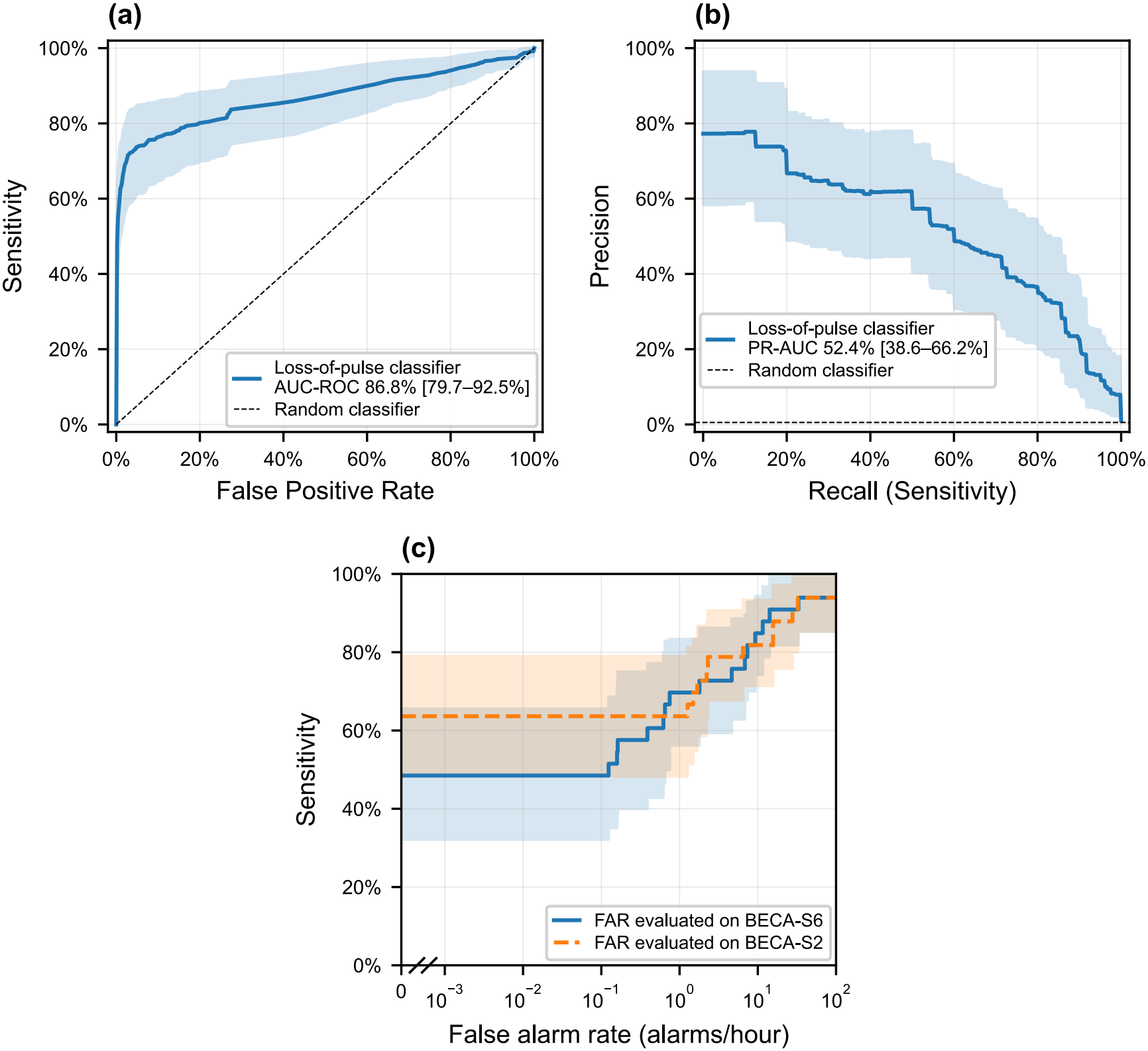
(a) The second-level Receiver Operating Characteristic curve. (b) The second-level Precision-Recall curve. (c) The event-level Free-response Receiver Operating Characteristic curves, with the false alarm rate measured on BECA-S6 or on BECA-S2 (see legend). All estimates are provided with 95% patient-cluster bootstrap confidence intervals.

At an event level, false alarm rates of 0.39 (95% CI: 0.00 to 4.64) and 7.36 (95% CI: 0.62 to 14.16) false alarms per hour are achieved at sensitivity operating points of 60% and 80%, respectively, measured over 156.7 wrist-hours of BECA-S6 data. The 95% sensitivity operating point was not reached within the evaluated range of up to 100 false alarms per hour (lower 95% confidence bound: 11.56 false alarms per hour). The false alarm rate remains zero up to a sensitivity of 48.5% (95% CI: 32.1 to 65.5%). When the false alarm rate was instead measured on BECA-S2, the sensitivity at a false alarm rate of zero was 63.6% (95% CI: 48.3 to 78.9%), and the false alarm rates at 60% and 80% sensitivity were 0.00 (95% CI: 0.00 to 2.22) and 6.47 (95% CI: 1.27 to 27.76) false alarms per hour; 95% sensitivity was not reached (lower 95% confidence bound: 15.67 false alarms per hour). The Free-response ROC curves are shown in Figure 5(c), and their tabular data are provided in Appendix C.

### 3.4 Subgroup analysis

Considering the effects of subgroups, we estimated the average marginal effects of the participant characteristics in both cohorts (Figure 6). In BECA-S2, 22 of the 31 patients had a defined per-patient PR-AUC (second-level PR-AUC is undefined for the 9 patients without recorded events), of whom 2 lacked a recorded Fitzpatrick skin tone; the model was therefore fit on the 20 patients with complete data. In BECA-S6, we modeled the false alarm count of all 100 participants.

**Figure 6:**
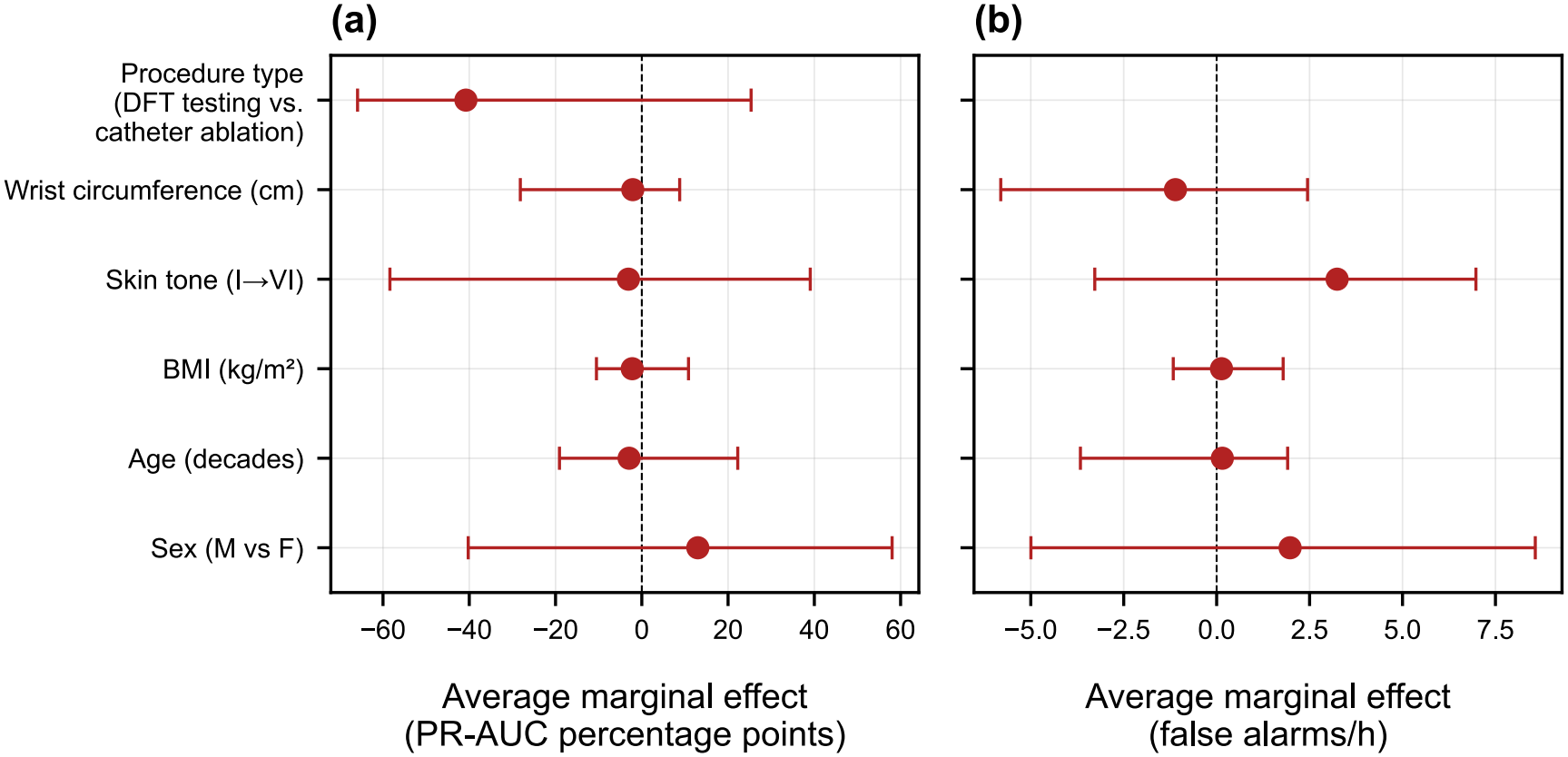
Average marginal effects of the participant characteristics on (a) the per-patient area under the precision-recall curve in BECA-S2 (*n* = 20, in percentage points) and (b) the false alarm rate in BECA-S6 (*n* = 100, in false alarms per hour). Error bars denote 95% percentile-bootstrap confidence intervals. None of the effects were significant before or after Bonferroni correction.

In both cohorts, none of the characteristics were associated with PR-AUC (BECA-S2) or the false alarm rate (BECA-S6) before or after Bonferroni correction (all *p*_Bonf_ = 1.0). In BECA-S2, all confidence intervals are wide, reflecting the small cohort and uneven subgroup sizes (e.g., 3 of 20 patients female) rather than well-estimated effects. Generalized variance inflation factors were low in both models (1.42–2.45 in BECA-S2 and 1.05–3.21 in BECA-S6), indicating that these results are not an artifact of collinearity among predictors. All average marginal effects and generalized variance inflation factors are provided in Appendix D.

## 4 Discussion

In this work, a model detected loss of pulse in wrist-PPG and accelerometry during routine cardiac procedures (BECA-S2) and event-free free-living conditions (BECA-S6) with a second-level AUC-ROC of 86.8% (95% CI: 79.7 to 92.5%) and PR-AUC of 52.4% (95% CI: 38.6 to 66.2%). At event level, with false alarms counted in free-living conditions, the false alarm rate remained zero up to a sensitivity of 48.5% (95% CI: 32.1 to 65.5%), after which it increased to 7.36 false alarms per hour at a sensitivity of 80%; the 95% sensitivity target was not reached within the evaluated false alarm rate range (up to 100 alarms per hour). In other words, about half of the loss-of-pulse events were detected without any false alarms in the roughly 157 wrist-hours of daily-life data, but higher sensitivities came at the cost of many false alarms.

While this work evaluates false alarm rate on a separate dataset capturing free-living conditions, counting false alarms on the BECA-S2 data captured during cardiac procedures instead gave a higher sensitivity at a false alarm rate of zero (63.6%, 95% CI: 48.3 to 78.9%, versus 48.5%) and slightly lower false alarm rates at 60% and 80% sensitivity (0.00 versus 0.39 and 6.47 versus 7.36 false alarms per hour), with overlapping confidence intervals throughout; neither reached 95% sensitivity. The difference at low false alarm rates could be explained by the state of the wearer: the free-living conditions in BECA-S6 may form a bigger challenge than the anesthetized and supine conditions in BECA-S2.

The DETECT studies used induced arrests in patients during cardiac procedures, including VT ablation and implantable defibrillator implantation, and validated their algorithm in independent patients. They reported sensitivities of 92–98% at 0.12–0.26 false alarms per hour [7, 8]. At comparable sensitivity (90%), our model produced more false alarms: 14.16 per hour (lower 95% confidence bound: 7.36). This may relate to the differences between the datasets and algorithms:

DETECT used a fixed rule-based algorithm developed on a considerably larger number of in-hospital patients (205 in DETECT-1), whereas our model was trained and evaluated on a combination of 31 in-hospital patients and 100 participants in free-living conditions.

The HEART-SAFE results (90.3% sensitivity at roughly 0.06 false alarms per hour) cannot be compared with ours, as their algorithm was developed and tested on the same data [9], so that overfitting to these data cannot be ruled out and their performance estimates may be optimistic. Google Research’s algorithm was developed and tested mainly outside the clinical setting, on healthy volunteers and stunt performers with tourniquet-induced peripheral pulselessness and on free-living data. It reported a sensitivity of 67.2% at 1.34 false alarms per user-year, equivalent to about 1.5 × 10^*−*4^ false alarms per hour [6]. This false alarm rate was based on 21.67 user-years of free-living data, whereas our 157 wrist-hours of BECA-S6 data cannot resolve false alarm rates with such resolution. However, given the need for an acceptably low false alarm rate, our data agree with Google’s choice of a low-sensitivity operating point.

A methodological point of wider relevance is target leakage. In studies where loss of pulse is induced or terminated by a procedure, such as cuff inflation, electrical VF induction, or defibrillation shocks, the procedure leaves signal artifacts that coincide with the label, and a model can exploit these artifacts instead of detecting the loss of pulse itself. In our study, not dealing with induction and shock artifacts as a form of target leakage would have increased the model’s scores: inserting a synthetic gap, which stands in for an induction or shock artifact, in the arrest context shifted the unmitigated model’s scores upward by 35.3% of its discriminative range, far outside the 5% tolerance. A gradient penalty on the model’s dependence on gapped input reduced this shift to − 1.23% (95% CI: − 3.46 to − 0.09%) at a penalty weight of 10^5^. While the PR-AUC point estimate was lower with the penalty (68.7% versus 52.4%), the paired difference (− 16.3 pp, 95% CI: − 34.1 to 1.8) was not statistically significant (*p* = 0.079). To our knowledge, this is the first study of smartwatch-based loss-of-pulse detection to mitigate target leakage. We recommend that future studies with induced or procedure-related events either avoid developing models on such data, as Google Research did with their electrophysiology-laboratory recordings of induced VF [6], or adopt both the synthetic-gap test, which can be applied to any trained model without retraining, and a mitigation strategy. Without mitigation, performance estimates risk being overly optimistic.

Exploratory subgroup analyses found no evidence that PR-AUC in BECA-S2 or the false alarm rate in BECA-S6 differed by participant characteristics. These analyses had limited power, in particular for sex and procedure type in the small BECA-S2 cohort and for the darker Fitzpatrick skin tones (IV–V, with no type VI), and therefore cannot establish equal performance across subgroups; larger and more diverse cohorts are needed.

### 4.1 Limitations

First, while false alarm rates were tested in out-of-hospital conditions, sensitivity was tested only on loss-of-pulse events that occurred in clinical settings during cardiac procedures, in patients who were typically sedated or anesthetized, lying supine, and largely motionless. Real OHCA occurs in free-living conditions with different posture, motion, and circulatory dynamics, so our results cannot be directly translated to real OHCA.

Second, sensitivity and false alarm rate were measured in cohorts with different demographics: the BECA-S6 cohort is relatively young and healthy (median age 31) compared to the BECA-S2 cohort with older patients (median age 60) who had cardiac disease.

Third, the BECA-S2 cohort is small (33 events in 22 patients), so all estimates have wide confidence intervals, and the statistical tests had limited power. This applies in particular to the target leakage test, which could only use the 13 events in 7 patients not preceded by a gap, and to the subgroup analysis, which included 20 patients, of whom only three were female. Similarly, darker skin types were underrepresented: only one BECA-S2 patient had a Fitzpatrick skin type darker than type II. Future studies should include more data, more female patients, and more participants with darker skin types to produce narrower confidence intervals.

Lastly, the loss-of-pulse events were shorter (median 10 s) than the loss of pulse in most OHCA cases, leading to shorter selected debounce lengths. Longer debounce lengths are likely to have led to fewer false alarms, making our performance estimations conservative. Future studies should include longer loss-of-pulse events to arrive at more realistic performance estimates.

## 5 Conclusion

We evaluated smartwatch-based detection of loss of pulse in cohorts of patients undergoing cardiac procedures and participants in free-living conditions. The model reached an AUC-ROC of 86.8% and a PR-AUC of 52.4%. About half of the loss-of-pulse events (48.5% sensitivity) were detected without any false alarms in daily-life data of healthy volunteers. Higher sensitivities came at the cost of more false alarms. Target leakage affected the model scores; a gradient penalty mitigated it, and we recommend that future studies test for target leakage and mitigate it. These results show the feasibility of smartwatch-based loss-of-pulse detection and provide a basis for larger studies with data more representative of real OHCA events.

## Author contributions

**Roelof G. Hup**: Conceptualization, Data curation, Formal analysis, Investigation, Methodology, Project administration, Resources, Software, Validation, Visualization, Writing – original draft. **Maud E. Kortman**: Investigation, Project administration, Writing – review & editing. **Robin W.F.H. Kuijpers**: Investigation, Project administration, Writing – review & editing. **Michelle J.S. Mensinck**: Investigation, Project administration, Writing – review & editing. **Chaimae Bouchnaf** : Writing – review & editing. **Lukas R.C. Dekker**: Conceptualization, Data curation, Funding acquisition, Methodology, Supervision, Writing – review & editing. **Alexandre J. Ouss**: Data curation, Writing – review & editing. **Hanno L. Tan**: Conceptualization, Funding acquisition, Writing – review & editing. **Reinder Haakma**: Conceptualization, Methodology, Resources, Supervision. **Xi Long**: Methodology, Supervision, Writing – review & editing. **Rik Vullings**: Conceptualization, Funding acquisition, Methodology, Project administration, Supervision, Writing – review & editing.

## Acknowledgments

During the preparation of this manuscript, the authors used Anthropic Claude to assist with code development and to support editing and refinement of the written text. All AI-generated output was reviewed, verified, and edited by the authors, who take full responsibility for the content, accuracy, and integrity of the manuscript.

## Data availability

Due to the sensitive nature of the data and constraints of the informed consent obtained, de-identified BECA-S2 and/or BECA-S6 data may be made available by the corresponding author upon reasonable request, subject to review of a research proposal to confirm compatibility with the scope of patient consent, and to a data sharing agreement. The code is available at https://github.com/gerbenhup/cardiac-arrest-detection.

