## Appendix A for "Smartwatch-based loss-of-pulse detection for out-of-hospital cardiac arrest: the BECA-S2 and BECA-S6 studies"

### A Feature overview

Table A.1 lists the 37 per-sample features extracted from the pulsatile photoplethysmogram (PPG) and accelerometer (ACC) signals, plus the binary gap-indicator feature appended afterward, for a total of 38 input channels per sample. All features are computed causally (using only current and past samples), after the accelerometry was downsampled to 32 Hz with a zero-phase anti-aliasing filter. Let $f_s$ denote the sampling frequency and $t$ a discrete sample index. The following notation is used throughout:

- $x_{\mathrm{dc}}(t)$: the low-pass filtered (baseline) PPG signal; $x_{\mathrm{ac}}(t)$: the high-pass filtered (pulsatile) PPG signal. Both use a third-order causal Butterworth filter with a 0.5 Hz cutoff (low-pass and high-pass, respectively).
- $a_k(t)$ for $k \in \{x, y, z\}$: the raw (gain-corrected) accelerometer axes; $\tilde{a}_k(t)$: the corresponding high-pass filtered (gravity-removed) axes, using a third-order causal Butterworth filter with a 0.5 Hz cutoff.
- For a signal $x$ and window length $N$ samples (ending at $t$, i.e. spanning $j \in \{t-N+1, \ldots, t\}$):

$$
\begin{aligned}
\bar{x}_N(t) &= \frac{1}{N} \sum_{j=t-N+1}^{t} x(j) \\
\mu_2[x](t) &= \frac{1}{N} \sum_{j=t-N+1}^{t} \left(x(j) - \bar{x}_N(t)\right)^2 \\
\mu_3[x](t) &= \frac{1}{N} \sum_{j=t-N+1}^{t} \left(x(j) - \bar{x}_N(t)\right)^3 \\
\mu_4[x](t) &= \frac{1}{N} \sum_{j=t-N+1}^{t} \left(x(j) - \bar{x}_N(t)\right)^4 \\
\mathrm{Var}_N[x](t) &= \mu_2[x](t) \\
\mathrm{Skew}_N[x](t) &= \mu_3[x](t) \,/\, \mu_2[x](t)^{3/2} \\
\mathrm{Kurt}_N[x](t) &= \mu_4[x](t) \,/\, \mu_2[x](t)^2 - 3
\end{aligned}
$$

- $v(t) = x_{\mathrm{ac}}(t) - x_{\mathrm{ac}}(t-1)$: the first derivative (velocity) of the pulsatile PPG signal; $p(t) = v(t) - v(t-1)$: its second derivative (acceleration).
- $\Delta_\tau[x](t) = \ln(x(t) + \varepsilon) - \ln(x(t - \tau f_s) + \varepsilon)$, with $\varepsilon = 10^{-6}$: the log-ratio of a quantity $x$ against its own value $\tau$ seconds earlier.

Time-domain window lengths are 2 s unless noted otherwise; frequency-domain features use an 8 s Hann-windowed FFT.

**Table A.1:** Overview of the 37 extracted features plus the gap-indicator feature (channel 37).

| Ch. | Feature | Description | Window |
|---|---|---|---|
| 0 | PPG baseline | $x_{\mathrm{dc}}(t)$ | – |
| 1 | PPG baseline derivative | $x_{\mathrm{dc}}(t) - x_{\mathrm{dc}}(t-1)$ | – |
| 2 | PPG pulsatile signal | $x_{\mathrm{ac}}(t)$ | – |
| 3 | PPG variance | $\ln(\mathrm{Var}_N[x_{\mathrm{ac}}](t) + \varepsilon)$ | 2 s |

*continued on next page*

| Ch. | Feature | Description | Window |
|---|---|---|---|
| 4 | PPG skewness | $\mathrm{Skew}_N[x_{\mathrm{ac}}](t)$ | 2 s |
| 5 | PPG kurtosis | $\mathrm{Kurt}_N[x_{\mathrm{ac}}](t)$ | 2 s |
| 6 | PPG pulsatility index | $\sqrt{\mathrm{Var}_N\left[x_{\mathrm{ac}}\right](t)} \,/\, \max\bigl(\lvert x_{\mathrm{dc}}(t)\rvert\,,\,10^{-3}\bigr)$ | 2 s |
| 7 | PPG velocity | $v(t) = x_{\mathrm{ac}}(t) - x_{\mathrm{ac}}(t-1)$ | – |
| 8 | PPG acceleration | $p(t) = v(t) - v(t-1)$ | – |
| 9 | Hjorth mobility | $\sqrt{\mathrm{Var}_N[v](t) \,/\, \mathrm{Var}_N[x_{\mathrm{ac}}](t)}$ | 2 s |
| 10 | Hjorth complexity | $\sqrt{\mathrm{Var}_N[p](t)\,\mathrm{Var}_N[x_{\mathrm{ac}}](t)} \,/\, \mathrm{Var}_N[v](t)$ | 2 s |
| 11 | PPG zero-crossing rate | $\dfrac{1}{2(N-1)} \displaystyle\sum_{j=t-N+1}^{t-1} \bigl\lvert\, \mathrm{sgn}(x_{\mathrm{ac}}(j{+}1)) - \mathrm{sgn}(x_{\mathrm{ac}}(j)) \,\bigr\rvert$ | 2 s |
| 12 | PPG peak-to-peak amplitude | $\ln(A(t) + \varepsilon)$ <br> $A(t) = \max_{j\in N} x_{\mathrm{ac}}(j) - \min_{j\in N} x_{\mathrm{ac}}(j)$ | 2 s |
| 13 | PPG amplitude log-ratio, 10 s | $\Delta_{10}[A](t)$ | 2 s + 10 s |
| 14 | PPG amplitude log-ratio, 30 s | $\Delta_{30}[A](t)$ | 2 s + 30 s |
| 15 | PPG amplitude log-ratio, 60 s | $\Delta_{60}[A](t)$ | 2 s + 60 s |
| 16 | PPG amplitude log-ratio, 120 s | $\Delta_{120}[A](t)$ | 2 s + 120 s |
| 17 | PPG signal energy | $\ln\left(\displaystyle\sum_{j=t-N+1}^{t} x_{\mathrm{ac}}(j)^2 + 1\right)$ | 2 s |
| 18 | PPG signal-to-noise ratio | $\displaystyle\sum_{f\in[0.5,4]\ \mathrm{Hz}} P(f,t) \Bigg/ \sum_{f\notin(0.5,5)\ \mathrm{Hz}} P(f,t)$ | 8 s |
| 19 | PPG spectral entropy | $-\displaystyle\sum_{f\in[0.5,4]\ \mathrm{Hz}} q(f,t)\ln q(f,t)$ <br> $q(f,t) = P(f,t)/\sum_{f'\in[0.5,4]\ \mathrm{Hz}} P(f',t)$ | 8 s |
| 20 | PPG spectral flux | $\ln\left(\dfrac{1}{n_f}\displaystyle\sum_{f}\bigl\lvert P(f,t) - P(f,t-1)\bigr\rvert + \varepsilon\right)$ | 8 s |
| 21 | Raw accelerometer, $x$-axis | $a_x(t)$ | – |
| 22 | Raw accelerometer, $y$-axis | $a_y(t)$ | – |

*continued on next page*

| Ch. | Feature | Description | Window |
|---|---|---|---|
| 23 | Raw accelerometer, $z$-axis | $a_z(t)$ | – |
| 24 | Filtered accelerometer, $x$-axis | $\tilde{a}_x(t)$ | – |
| 25 | Filtered accelerometer, $y$-axis | $\tilde{a}_y(t)$ | – |
| 26 | Filtered accelerometer, $z$-axis | $\tilde{a}_z(t)$ | – |
| 27 | ACC log range-sum | $\ln(R(t)+1)$ <br> $R(t) = \sum_{k\in\{x,y,z\}} \left[\max_{j\in N} \tilde{a}_k(j) - \min_{j\in N} \tilde{a}_k(j)\right]$ | 2 s |
| 28 | ACC range-sum log-ratio, 10 s | $\Delta_{10}[R](t)$ | 2 s + 10 s |
| 29 | ACC range-sum log-ratio, 30 s | $\Delta_{30}[R](t)$ | 2 s + 30 s |
| 30 | ACC range-sum log-ratio, 60 s | $\Delta_{60}[R](t)$ | 2 s + 60 s |
| 31 | ACC range-sum log-ratio, 120 s | $\Delta_{120}[R](t)$ | 2 s + 120 s |
| 32 | ACC variance | $\ln\left(\sqrt{\sum_{k\in\{x,y,z\}} \mathrm{Var}_N[\tilde{a}_k](t)^2} + \varepsilon\right)$ | 2 s |
| 33 | ACC skewness | $\sqrt{\sum_{k\in\{x,y,z\}} \mathrm{Skew}_N[\tilde{a}_k](t)^2}$ | 2 s |
| 34 | ACC kurtosis | $\sqrt{\sum_{k\in\{x,y,z\}} \mathrm{Kurt}_N[\tilde{a}_k](t)^2}$ | 2 s |
| 35 | Acceleration magnitude | $\sqrt{\sum_{k\in\{x,y,z\}} a_k(t)^2}$ | – |
| 36 | Jerk magnitude | $\ln\left(\sqrt{\sum_{k\in\{x,y,z\}} (a_k(t) - a_k(t-1))^2} + \varepsilon\right)$ | – |
| 37 | Gap indicator | $g(t) \in \{0,1\}$: 1 if sample $t$ lies in a gapped second; 0 otherwise. | – |
