## Appendix B for "Smartwatch-based loss-of-pulse detection for out-of-hospital cardiac arrest: the BECA-S2 and BECA-S6 studies"

### B Network architecture

The network consists of a per-second encoder and a sequence decoder. The encoder (Table B.2) maps each second's local feature window, of size $32 \times 38$, to a single embedding vector; the same encoder (shared weights) is applied independently to every second in the input sequence. An LSTM processes these vector embeddings into context-aware vector embeddings. The decoder (Table B.3) then produces one logit per second. The full network combines encoder, LSTM, and decoder (Table B.4). The hyperparameter space is given in Table B.1.

**Table B.1:** Hyperparameter search space. The gradient-penalty weight was selected with the target-leakage tests (Section 2.5) and the debounce length per fold after model selection; neither was part of the per-fold successive-halving search.

| **Hyperparameter** | **Search space** |
|---|---|
| Model width ($f$) | $\{8, 16, 32\}$ |
| Encoder dropout rate ($d$) | $\{0.2, 0.3, 0.4, 0.5, 0.6, 0.7\}$ |
| Tversky weights ($\alpha, \beta$) | $\{(0.7, 0.3), (0.6, 0.4), (0.5, 0.5), (0.4, 0.6), (0.3, 0.7)\}$ |
| Smoothness-penalty weight ($\lambda$) | $\{0.0, 0.1, 0.2\}$ |
| Gradient-penalty weight ($w$) | $\{0, 1, 10, 10^2, 10^3, 10^4, 10^5, 10^6, 10^7\}$ |
| Debounce length | [1, 60] |

**Table B.2:** Per-second encoder. Applied independently to each of the $T$ seconds in the input sequence.

| **Layer** | **Output size** |
|---|---|
| Input | $32 \times 38$ |
| Convolution ($f$ filters, kernel size 3) + ReLU | $32 \times f$ |
| Batch normalization | $32 \times f$ |
| Dropout (rate $d$) | $32 \times f$ |
| Max pooling (kernel size 2) | $16 \times f$ |
| Convolution ($2f$ filters, kernel size 3) + ReLU | $16 \times 2f$ |
| Batch normalization | $16 \times 2f$ |
| Dropout (rate $d$) | $16 \times 2f$ |
| Max pooling (kernel size 2) | $8 \times 2f$ |
| Convolution ($4f$ filters, kernel size 3) + ReLU | $8 \times 4f$ |
| Batch normalization | $8 \times 4f$ |
| Global average pooling | $4f$ |

**Table B.3:** Per-second decoder. Consumes a context-aware per-second embedding from the LSTM.

| **Layer** | **Output size** |
|---|---|
| Input | $8f$ |
| Fully connected (1 unit) | 1 |

**Table B.4:** The complete network.

| **Stage** | **Output size** |
|---|---|
| Input | $T \times 32 \times 38$ |
| Time-distributed encoder (Table B.2) | $T \times 4f$ |
| LSTM ($8f$ units) | $T \times 8f$ |
| Time-distributed decoder (Table B.3) | $T \times 1$ |
