## Appendix C for "Smartwatch-based loss-of-pulse detection for out-of-hospital cardiac arrest: the BECA-S2 and BECA-S6 studies"

### C FROC tabular data

Table C.1 lists the event-level false alarm rate (false alarms per hour) required to reach each sensitivity, with false alarms counted on BECA-S6 and on BECA-S2.

**Table C.1:** Event-level false alarm rate (false alarms per hour) required to reach each sensitivity of the BECA-S2 events, counted on BECA-S6 (left) or on BECA-S2 (right), gradient-penalty weight $w = 10^5$. Point estimate from the micro-averaged FROC curve, with 95% percentile confidence intervals from a patient-cluster bootstrap (2000 resamples). n.r.: not reached within the evaluated range (up to 100 false alarms per hour); resamples that do not reach a sensitivity level are counted as exceeding every false alarm rate.

| | **False alarms counted on BECA-S6** | | **False alarms counted on BECA-S2** | |
|---|---|---|---|---|
| **Sensitivity (%)** | **False alarms per hour** | **95% CI** | **False alarms per hour** | **95% CI** |
| 0 | 0.00 | 0.00–0.00 | 0.00 | 0.00–0.00 |
| 5 | 0.00 | 0.00–0.00 | 0.00 | 0.00–0.00 |
| 10 | 0.00 | 0.00–0.00 | 0.00 | 0.00–0.00 |
| 15 | 0.00 | 0.00–0.00 | 0.00 | 0.00–0.00 |
| 20 | 0.00 | 0.00–0.00 | 0.00 | 0.00–0.00 |
| 25 | 0.00 | 0.00–0.00 | 0.00 | 0.00–0.00 |
| 30 | 0.00 | 0.00–0.00 | 0.00 | 0.00–0.00 |
| 35 | 0.00 | 0.00–0.12 | 0.00 | 0.00–0.00 |
| 40 | 0.00 | 0.00–0.16 | 0.00 | 0.00–0.00 |
| 45 | 0.00 | 0.00–0.62 | 0.00 | 0.00–0.00 |
| 50 | 0.12 | 0.00–0.65 | 0.00 | 0.00–1.27 |
| 55 | 0.16 | 0.00–0.75 | 0.00 | 0.00–1.68 |
| 60 | 0.39 | 0.00–4.64 | 0.00 | 0.00–2.22 |
| 65 | 0.65 | 0.00–6.84 | 1.27 | 0.00–2.30 |
| 70 | 1.80 | 0.16–9.27 | 1.68 | 0.00–6.47 |
| 75 | 4.64 | 0.16–11.56 | 2.22 | 0.00–15.67 |
| 80 | 7.36 | 0.62–14.16 | 6.47 | 1.27–27.76 |
| 85 | 11.56 | 1.80–33.08 | 15.67 | 1.68–32.47 |
| 90 | 14.16 | 7.36–n.r. | 27.76 | 2.30–n.r. |
| 95 | n.r. | 11.56–n.r. | n.r. | 15.67–n.r. |
| 100 | n.r. | 14.16–n.r. | n.r. | 27.76–n.r. |
