## Appendix D for "Smartwatch-based loss-of-pulse detection for out-of-hospital cardiac arrest: the BECA-S2 and BECA-S6 studies"

### D Patient and participant subgroup analysis

Table D.1 provides the average marginal effects (in percentage points) and generalized variance inflation factors (GVIF) for the subgroup analysis of per-patient PR-AUC in BECA-S2. Table D.2 provides the average marginal effects (in false alarms per hour) and GVIF for the subgroup analysis of the false alarm rate in BECA-S6.

**Table D.1:** Fractional-logit regression of per-patient PR-AUC on patient characteristics in BECA-S2 (gradient-penalty weight $w = 10^5$, $n = 20$): average marginal effects (AME) in percentage points with 95% confidence intervals, raw and Bonferroni-corrected $p$-values, and generalized variance inflation factors (GVIF).

| **Predictor** | **AME, pp (95% CI)** | $p$ **(raw)** | $p$ **(Bonferroni)** | **GVIF** |
|---|---|---|---|---|
| Sex (male vs. female) | 12.99 (−40.24, 58.03) | 0.672 | 1.000 | 1.42 |
| Age (per decade) | −2.94 (−19.09, 22.25) | 0.851 | 1.000 | 2.03 |
| BMI (kg/m$^2$) | −2.21 (−10.51, 10.83) | 0.759 | 1.000 | 2.45 |
| Skin tone (Fitzpatrick I–VI) | −3.10 (−58.40, 39.06) | 0.965 | 1.000 | 1.70 |
| Wrist circumference (cm) | −2.09 (−28.16, 8.78) | 0.617 | 1.000 | 1.91 |
| Procedure type (DFT testing vs. catheter ablation) | −40.78 (−65.90, 25.34) | 0.249 | 1.000 | 2.35 |

**Table D.2:** Poisson regression of the false alarm count on participant characteristics in BECA-S6 ($n = 100$, both wrists summed, log monitored hours as offset): average marginal effects (AME) in false alarms per hour with 95% bootstrap confidence intervals, raw and Bonferroni-corrected $p$-values, and generalized variance inflation factors (GVIF).

| **Predictor** | **AME, FA/h (95% CI)** | $p$ **(raw)** | $p$ **(Bonferroni)** | **GVIF** |
|---|---|---|---|---|
| Sex (male vs. female) | 1.98 (−5.00, 8.57) | 0.570 | 1.000 | 1.53 |
| Age (per decade) | 0.15 (−3.66, 1.91) | 0.831 | 1.000 | 1.35 |
| BMI (kg/m$^2$) | 0.13 (−1.17, 1.79) | 0.727 | 1.000 | 2.35 |
| Skin tone (Fitzpatrick I–VI) | 3.24 (−3.28, 6.97) | 0.241 | 1.000 | 1.05 |
| Wrist circumference (cm) | −1.11 (−5.81, 2.45) | 0.544 | 1.000 | 3.21 |
